# Milder disease trajectory among COVID-19 patients hospitalised with the SARS-CoV-2 Omicron variant compared with the Delta variant in Norway

**DOI:** 10.1101/2022.03.10.22272196

**Authors:** Jeanette Stålcrantz, Anja Bråthen Kristoffersen, Håkon Bøås, Lamprini Veneti, Elina Seppälä, Nina Aasand, Olav Hungnes, Reidar Kvåle, Karoline Bragstad, Eirik Alnes Buanes, Robert Whittaker

## Abstract

Using individual-level national registry data, we conducted a cohort study to estimate differences in the length of hospital stay, and risk of admission to an intensive care unit and in-hospital death among patients infected with the SARS-CoV-2 Omicron variant, compared to patients infected with Delta variant in Norway. We included 409 (38%) patients infected with Omicron and 666 (62%) infected with Delta who were hospitalised with COVID-19 as the main cause of hospitalisation between 6 December 2021 and 6 February 2022. Omicron patients had a 48% lower risk of intensive care admission (aHR: 0.52, 95%CI: 0.34–0.80) and a 56% lower risk of in-hospital death (aHR: 0.44, 95%CI: 0.24–0.79) compared to Delta patients. Omicron patients had a shorter length of stay (with or without ICU stay) compared to Delta patients in the age groups from 18–79 years and those who had at least completed their primary vaccination. This supports growing evidence of reduced disease severity among hospitalised Omicron patients compared with Delta patients.

## Introduction

The first coronavirus disease (COVID-19) cases infected with the severe acute respiratory syndrome coronavirus 2 (SARS-CoV-2) Omicron variant (Phylogenetic Assignment of Named Global Outbreak Lineages (Pangolin) designation B.1.1.529) were detected in Norway on 26 November 2021 (1). By late December, Omicron had superseded the Delta variant (Pangolin designation B.1.617.2) as the dominant circulating variant (2).

In order to provide timely and ongoing support for patient management and capacity planning in hospitals in Norway, we estimated the length of hospital stay (LoS) and risk of admission to an intensive care unit (ICU) and in-hospital death among hospitalised patients infected with Omicron, compared to patients infected with Delta.

## Methods

### Study population

We conducted a cohort study on patients hospitalised with a positive SARS-CoV-2 test between 6 December 2021 and 6 February 2022. We included patients for whom COVID-19 was reported as the main cause of hospitalisation, who were known to be infected with the Omicron or Delta variant and who had a national identity number registered.

### Data sources

We extracted data from the Norwegian national emergency preparedness registry – Beredt C19 (3). This registry contains individual-level national data on all COVID-19 related hospitalisations, ICU admissions and deaths. Further details on the data sources and definitions, including the categorisation of variants, can be found in supplement A, section 1. Data were extracted on 15 February 2022, allowing a minimum 8 days of follow-up since last date of hospitalisation.

### Data analysis

Full details on the data analysis are presented in supplementary material A, section 2. Briefly, our outcomes were discharge from hospital (with and without ICU stay), admission to ICU, in-hospital death and a composite measure of ICU admission or in-hospital death. LoS was calculated as the time between first admission and last discharge. To estimate differences in our outcomes we calculated adjusted hazard ratios (aHR) using a Cox proportional hazards model. Explanatory variables included virus variant, sex, age, country of birth, underlying risk factors, regional health authority and vaccination status (see table 1). We also conducted subgroup analysis by age group and vaccination status for subgroups with ≥50 omicron and ≥50 delta patients, and ≥10 outcomes. Estimates from the univariable model for all patients and all multivariable models are presented in supplement B.

**Table 1.**
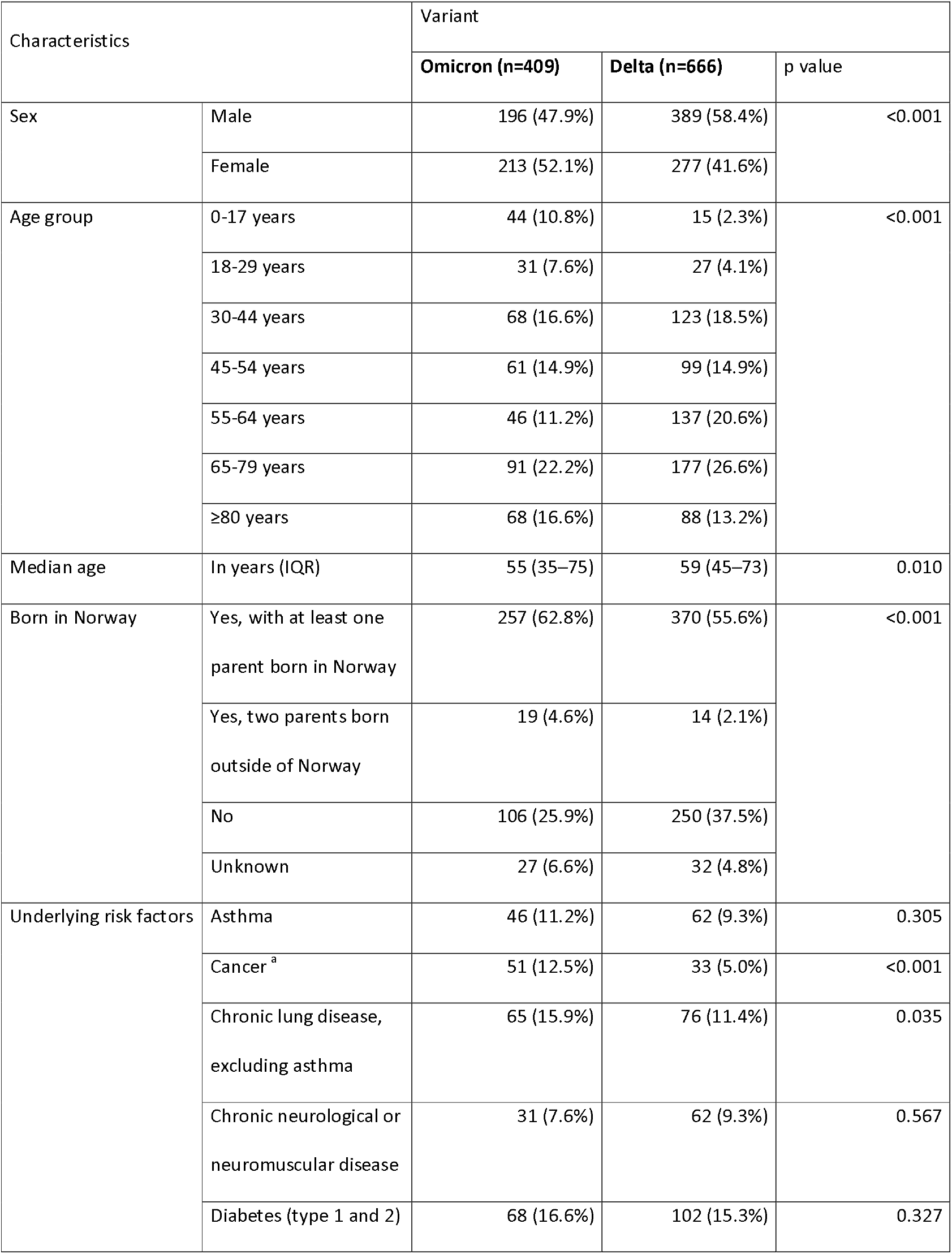
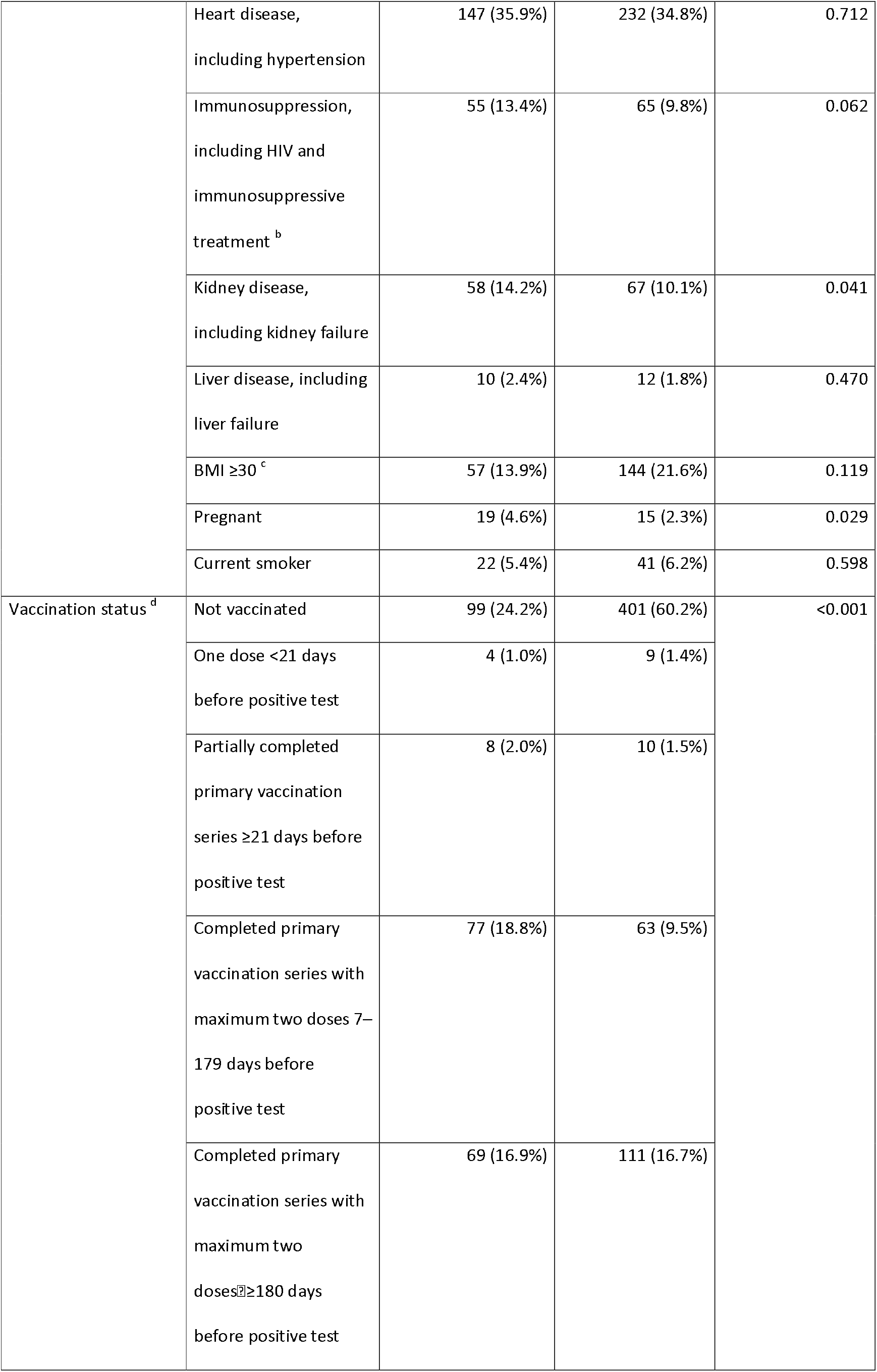
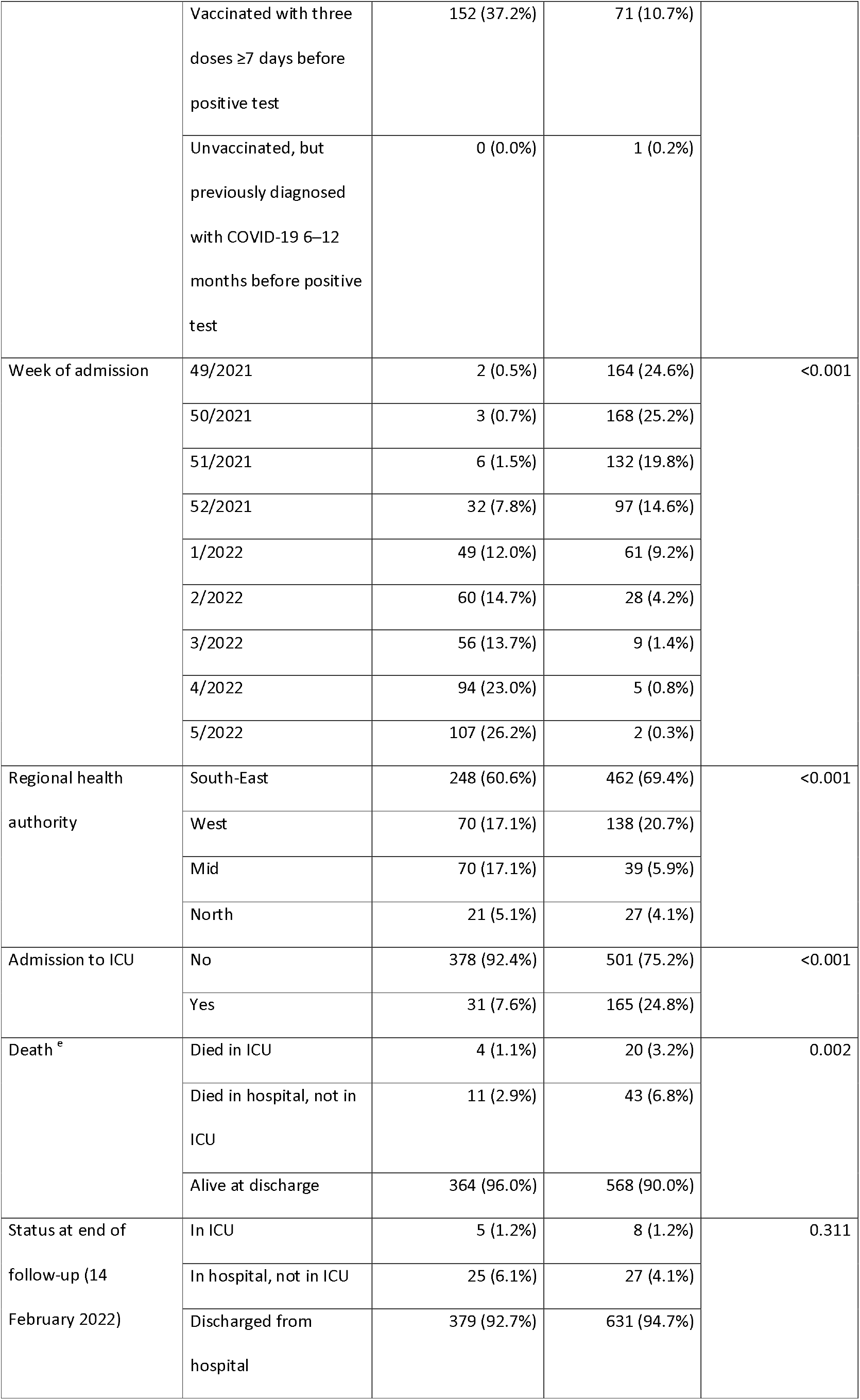
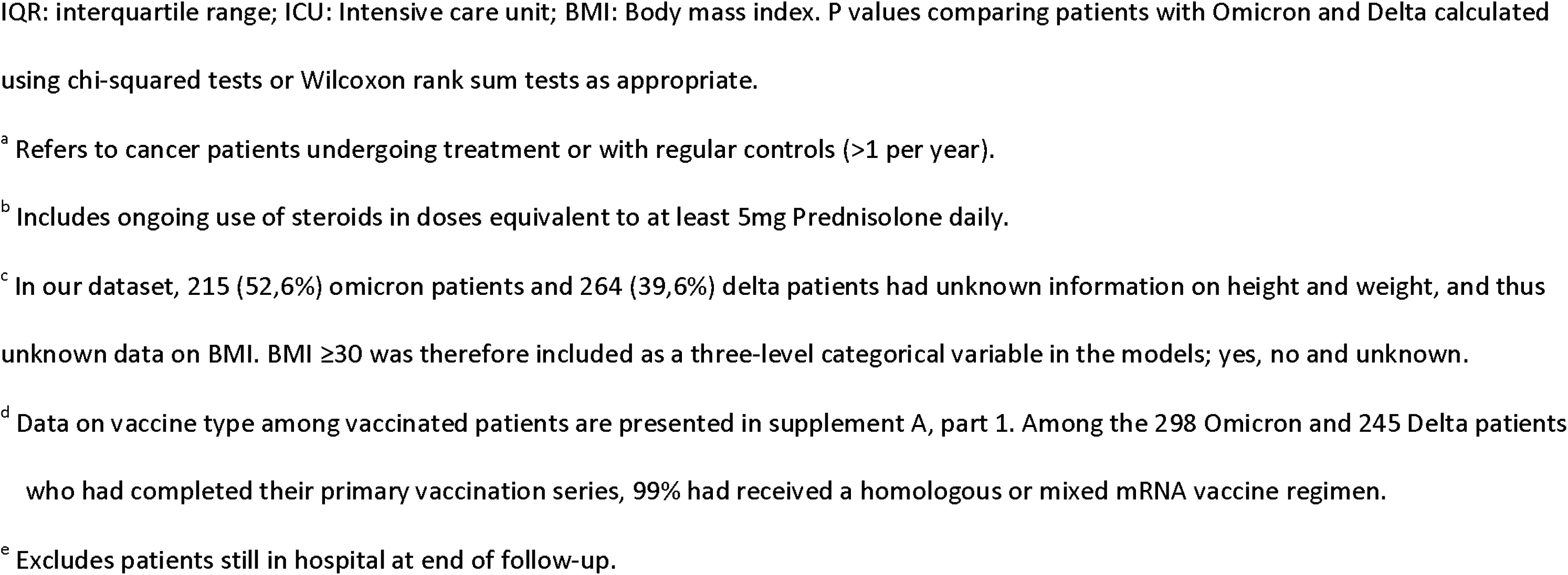
Characteristics of patients hospitalised with COVID-19 as the main cause of hospitalisation and infected with the SASRS-CoV-2 omicron or delta variant, by variant, Norway, 6 December 2021 – 6 February 2022 (n=1075)

| Characteristics |  | Variant |  |  |
| --- | --- | --- | --- | --- |
|  |  | Omicron (n=409) | Delta (n=666) | p value |
| Sex | Male | 196 (47.9%) | 389 (58.4%) | <0.001 |
|  | Female | 213 (52.1%) | 277 (41.6%) |  |
| Age group | 0-17 years | 44 (10.8%) | 15 (2.3%) | <0.001 |
|  | 18-29 years | 31 (7.6%) | 27 (4.1%) |  |
|  | 30-44 years | 68 (16.6%) | 123 (18.5%) |  |
|  | 45-54 years | 61 (14.9%) | 99 (14.9%) |  |
|  | 55-64 years | 46 (11.2%) | 137 (20.6%) |  |
|  | 65-79 years | 91 (22.2%) | 177 (26.6%) |  |
|  | ≥80 years | 68 (16.6%) | 88 (13.2%) |  |
| Median age | In years (IQR) | 55 (35–75) | 59 (45–73) | 0.010 |
| Born in Norway | Yes, with at least one parent born in Norway | 257 (62.8%) | 370 (55.6%) | <0.001 |
|  | Yes, two parents born outside of Norway | 19 (4.6%) | 14 (2.1%) |  |
|  | No | 106 (25.9%) | 250 (37.5%) |  |
|  | Unknown | 27 (6.6%) | 32 (4.8%) |  |
| Underlying risk factors | Asthma | 46 (11.2%) | 62 (9.3%) | 0.305 |
|  | Cancer <sup>a</sup> | 51 (12.5%) | 33 (5.0%) | <0.001 |
|  | Chronic lung disease, excluding asthma | 65 (15.9%) | 76 (11.4%) | 0.035 |
|  | Chronic neurological or neuromuscular disease | 31 (7.6%) | 62 (9.3%) | 0.567 |
|  | Diabetes (type 1 and 2) | 68 (16.6%) | 102 (15.3%) | 0.327 |
|  | Heart disease, including hypertension | 147 (35.9%) | 232 (34.8%) | 0.712 |
|  | Immunosuppression, including HIV and immunosuppressive treatment <sup>b</sup> | 55 (13.4%) | 65 (9.8%) | 0.062 |
|  | Kidney disease, including kidney failure | 58 (14.2%) | 67 (10.1%) | 0.041 |
|  | Liver disease, including liver failure | 10 (2.4%) | 12 (1.8%) | 0.470 |
| | BMI $\geq 30$ <sup>c</sup> | 57 (13.9%) | 144 (21.6%) | 0.119 |
|  | Pregnant | 19 (4.6%) | 15 (2.3%) | 0.029 |
|  | Current smoker | 22 (5.4%) | 41 (6.2%) | 0.598 |
| Vaccination status <sup>d</sup> | Not vaccinated | 99 (24.2%) | 401 (60.2%) | <0.001 |
|  | One dose <21 days before positive test | 4 (1.0%) | 9 (1.4%) |  |
| | Partially completed primary vaccination series $\geq 21$ days before positive test | 8 (2.0%) | 10 (1.5%) | |
|  | Completed primary vaccination series with maximum two doses 7–179 days before positive test | 77 (18.8%) | 63 (9.5%) |  |
| | Completed primary vaccination series with maximum two doses $\geq 180$ days before positive test | 69 (16.9%) | 111 (16.7%) | |
| | Vaccinated with three doses $\geq 7$ days before positive test | 152 (37.2%) | 71 (10.7%) | |
|  | Unvaccinated, but previously diagnosed with COVID-19 6–12 months before positive test | 0 (0.0%) | 1 (0.2%) |  |
| Week of admission | 49/2021 | 2 (0.5%) | 164 (24.6%) | <0.001 |
|  | 50/2021 | 3 (0.7%) | 168 (25.2%) |  |
|  | 51/2021 | 6 (1.5%) | 132 (19.8%) |  |
|  | 52/2021 | 32 (7.8%) | 97 (14.6%) |  |
|  | 1/2022 | 49 (12.0%) | 61 (9.2%) |  |
|  | 2/2022 | 60 (14.7%) | 28 (4.2%) |  |
|  | 3/2022 | 56 (13.7%) | 9 (1.4%) |  |
|  | 4/2022 | 94 (23.0%) | 5 (0.8%) |  |
|  | 5/2022 | 107 (26.2%) | 2 (0.3%) |  |
| Regional health authority | South-East | 248 (60.6%) | 462 (69.4%) | <0.001 |
|  | West | 70 (17.1%) | 138 (20.7%) |  |
|  | Mid | 70 (17.1%) | 39 (5.9%) |  |
|  | North | 21 (5.1%) | 27 (4.1%) |  |
| Admission to ICU | No | 378 (92.4%) | 501 (75.2%) | <0.001 |
|  | Yes | 31 (7.6%) | 165 (24.8%) |  |
| Death <sup>e</sup> | Died in ICU | 4 (1.1%) | 20 (3.2%) | 0.002 |
|  | Died in hospital, not in ICU | 11 (2.9%) | 43 (6.8%) |  |
|  | Alive at discharge | 364 (96.0%) | 568 (90.0%) |  |
| Status at end of follow-up (14 February 2022) | In ICU | 5 (1.2%) | 8 (1.2%) | 0.311 |
|  | In hospital, not in ICU | 25 (6.1%) | 27 (4.1%) |  |
|  | Discharged from hospital | 379 (92.7%) | 631 (94.7%) |  |

IQR: interquartile range; ICU: Intensive care unit; BMI: Body mass index. P values comparing patients with Omicron and Delta calculated using chi-squared tests or Wilcoxon rank sum tests as appropriate.
<sup>a</sup> Refers to cancer patients undergoing treatment or with regular controls (>1 per year).
<sup>b</sup> Includes ongoing use of steroids in doses equivalent to at least 5mg Prednisolone daily.
<sup>c</sup> In our dataset, 215 (52,6%) omicron patients and 264 (39,6%) delta patients had unknown information on height and weight, and thus unknown data on BMI. BMI $\geq 30$ was therefore included as a three-level categorical variable in the models; yes, no and unknown.
<sup>d</sup> Data on vaccine type among vaccinated patients are presented in supplement A, part 1. Among the 298 Omicron and 245 Delta patients who had completed their primary vaccination series, 99% had received a homologous or mixed mRNA vaccine regimen.
<sup>e</sup> Excludes patients still in hospital at end of follow-up.

We also assessed the representativeness of patients with known variant among all COVID-19 patients in the study period (supplementary material A, section 3).

### Ethics

Ethical approval was granted by Regional Committees for Medical Research Ethics - Southeast Norway, reference number 249509.

## Results

During the study period, 1747 patients were hospitalised with COVID-19 as the main cause of hospitalisation. Of these, 1710 (98%) had a national identity number, of which 1079 (63%) had known variant. Our study cohort comprised 409 Omicron (38%) and 666 Delta patients (62%). We excluded three patients reported to be infected with another variant, and one Delta patient with a date of positive test two months before hospitalisation. The median number of days from positive test to admission was 0 (interquartile range (IQR): 0–1) for Omicron patients and 4 (IQR: 0–7) for Delta patients. Detailed characteristics of the study cohort are presented in table 1. Descriptive results for each outcome by subgroup are presented in table 2.

**Table 2.** Number of patients, median number of days from admission to discharge from hospital, admissions to ICU and deaths in hospital among patients admitted to hospital with COVID-19 as the main cause of hospitalisation, by variant and subgroup, Norway, 6 December 2021 – 6 February 2022

|  | Omicron |  |  |  |  | Delta |  |  |  |  |
| --- | --- | --- | --- | --- | --- | --- | --- | --- | --- | --- |
|  | Number of patients | Median number of days from admission to discharge from hospital (IQR)<sup>a</sup> | Median number of days from admission to discharge, patients not admitted to ICU (IQR)<sup>a</sup> | Number admitted to ICU (%) | Number of deaths in hospital (%)<sup>b</sup> | Number of patients | Median number of days from admission to discharge from hospital (IQR)<sup>a</sup> | Median number of days from admission to discharge, patients not admitted to ICU (IQR)<sup>a</sup> | Number admitted to ICU (%) | Number of deaths in hospital (%)<sup>b</sup> |
| **All patients** | 409 | 2.82<br>(1.49–6.24) | 2.64<br>(1.30–4.94) | 31<br>(7.6%) | 15<br>(4.0%) | 666 | 5.93<br>(3.01–11.98) | 4.61<br>(2.50–7.77) | 165<br>(24.8%) | 63<br>(10.0%) |
| **Age groups** |  |  |  |  |  |  |  |  |  |  |
| – <18 years | 44 | <sup>c</sup> | <sup>c</sup> | <sup>c</sup> | <sup>c</sup> | 15 | <sup>c</sup> | <sup>c</sup> | <sup>c</sup> | <sup>c</sup> |
| – 18-44 years | 99 | 1.81<br>(0.83–3.00) | 1.80<br>(0.82–2.92) | 4<br>(4.0%) | 0<br>(0%) | 150 | 4.68<br>(2.67–8.12) | 3.61<br>(1.93–5.90) | 27<br>(18.0%) | 0<br>(0%) |
| – 45-64 years | 107 | 2.79<br>(1.67–7.64) | 2.63<br>(1.53–6.69) | 9<br>(8.4%) | 3<br>(3.1%) | 236 | 7.22<br>(3.27–15.10) | 4.61<br>(2.50–8.00) | 78<br>(33.1%) | 5<br>(2.3%) |
| – 65-79 years | 91 | 4.16<br>(2.10–7.84) | 3.97<br>(2.06–6.52) | 11<br>(12,1%) | 5<br>(6.2%) | 177 | 7.42<br>(3.88–13.65) | 6.08<br>(3.15–9.76) | 48<br>(27.1%) | 21<br>(12.6%) |
| – ≥80 years | 68 | 3.82<br>(2.05–7.66) | 3.82<br>(2.05–8.08) | 3<br>(4.4%) | 7<br>(11.1%) | 88 | 5.24<br>(2.76–8.31) | 4.56<br>(2.72–7.77) | 11<br>(12.5%) | 36<br>(40.9%) |
| **Vaccination status** |  |  |  |  |  |  |  |  |  |  |
| – Unvaccinated | 99 | 2.27<br>(0.88–4.31) | 2.13<br>(0.83–3.71) | 8<br>(8.1%) | 3<br>(3.2%) | 401 | 6.68<br>(3.15–12.35) | 4.87<br>(2.76–7.88) | 117<br>(29.2%) | 32<br>(8.4%) |
| – One dose <21 days before positive test | 4 | c | c | c | c | 9 | c | c | c | c |
| – Partially completed primary vaccination series ≥21 days before positive test | 8 | c | c | c | c | 10 | c | c | c | c |
| – Completed primary vaccination series with maximum two doses 7–179 days before positive test | 77 | 1.88<br>(0.97–2.92) | 1.83<br>(0.92–2.72) | 6<br>(7.6%) | 1<br>(1.4%) | 63 | 4.40<br>(1.38–6.93) | 3.16<br>(1.21–6.08) | 11<br>(17.5%) | 3<br>(5.2%) |
| – Completed primary vaccination series with maximum two doses ≥180 days before positive test | 69 | 3.82<br>(2.06–6.94) | 3.06<br>(1.86–6.07) | 9<br>(13.0%) | 3<br>(4.6%) | 111 | 4.88<br>(2.71–10.80) | 4.00<br>(2.09–7.77) | 15<br>(13.5%) | 17<br>(16.0%) |
| – Vaccinated with three doses ≥7 days before positive test | 152 | 3.79<br>(1.90–7.57) | 3.61<br>(1.86–7.12) | 7<br>(4.6%) | 8<br>(5.8%) | 71 | 6.94<br>(3.20–13.22) | 5.24<br>(2.56–9.76) | 18<br>(25.4%) | 10<br>(14.9%) |
| — Unvaccinated, but previously<br>diagnosed with COVID-19 6–12<br>months before positive test | 0 | <sup>c</sup> | <sup>c</sup> | <sup>c</sup> | <sup>c</sup> | 1 | <sup>c</sup> | <sup>c</sup> | <sup>c</sup> | <sup>c</sup> |

IQR: interquartile range; ICU: intensive care unit.
<sup>a</sup> Estimates from univariable Cox regression.
<sup>b</sup> Crude proportions calculated excluding those still admitted at the end of the study period. All patients: 30 Omicron, 35 Delta; 18–44 years: 2 Omicron, 6 Delta; 45–64 years: 11 Omicron, 19 Delta; 65–79 years: 10 Omicron, 10 Delta; ≥80 years: 5 Omicron, 0 Delta; Unvaccinated: 6 Omicron, 21 Delta; Completed primary vaccination series with maximum two doses 7–179 days before positive test: 4 Omicron, 5 Delta; Completed primary vaccination series with maximum two doses ≥180 days before positive test: 4 Omicron, 5 Delta; Vaccinated with three doses ≥7 days before positive test: 15 Omicron, 4 Delta.
<sup>c</sup> Subgroup analysis not conducted due to small sample size (<50 omicron and/or <50 delta patients and/or <10 outcomes).

Omicron patients had a 48% lower risk of ICU admission (aHR: 0.52, 95%CI: 0.34–0.80) and a 56% lower risk of in-hospital death (aHR: 0.44, 95%CI: 0.24–0.79), compared to Delta patients. By age subgroup, Omicron patients 18–79 years had a lower risk of ICU admission than Delta patients. We did not observe a difference in the risk of death between Omicron and Delta patients 65–79 years old. Patients ≥80 years were infrequently admitted to ICU, but Omicron patients had a lower risk of death than Delta patients. Omicron patients vaccinated with three doses had an 80% lower risk of ICU admission (aHR: 0.20, 95%CI: 0.08–0.47), and a 70% lower risk of in-hospital death (aHR: 0.30, 95%CI: 0.11–0.83). Results tended in the same direction for unvaccinated patients (aHR for ICU admission or death: 0.51, 95%CI: 0.26–0.98). We did not observe a difference in the risk of ICU admission or death between Omicron and Delta patients who had completed primary vaccination with maximum two doses (table 3).

**Table 3.**
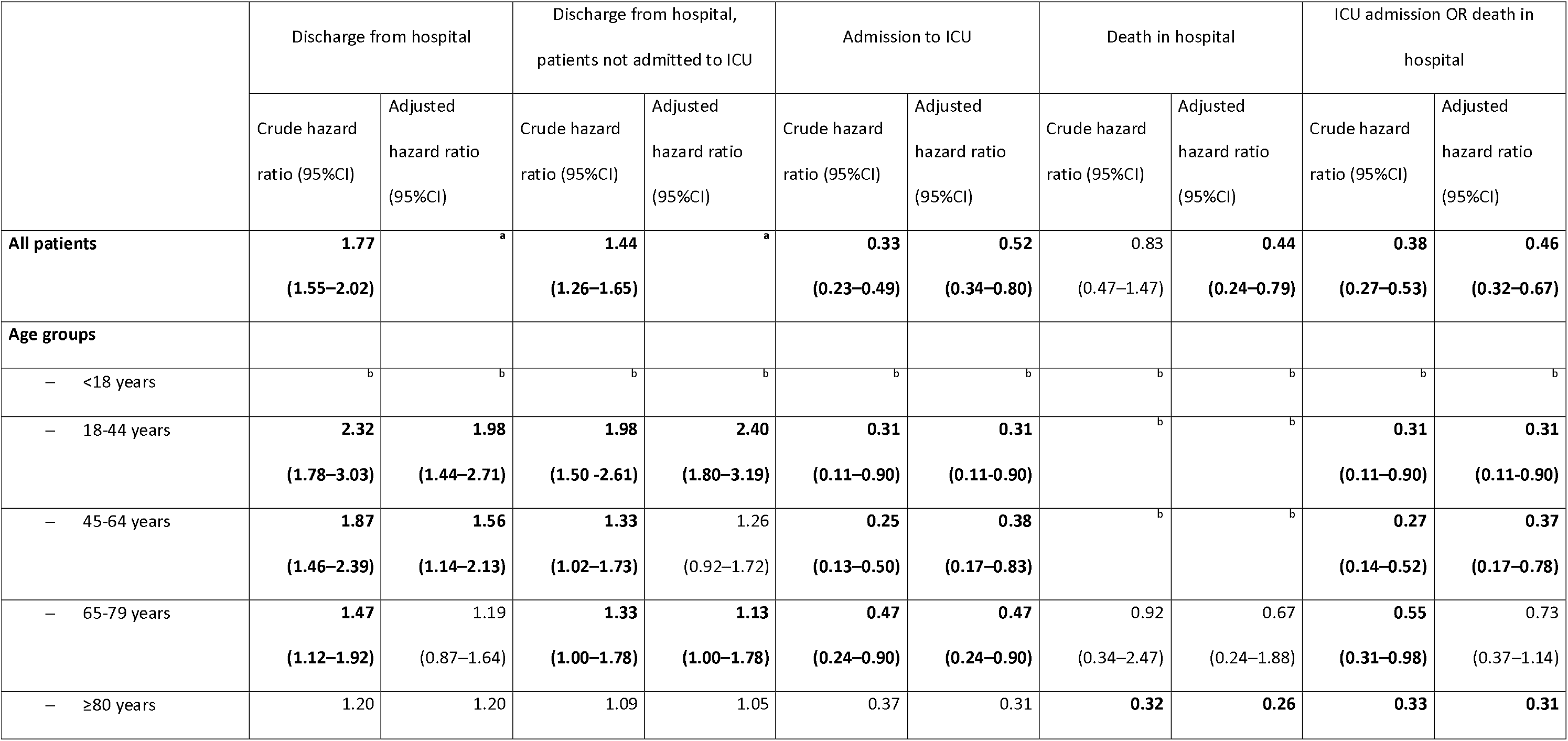
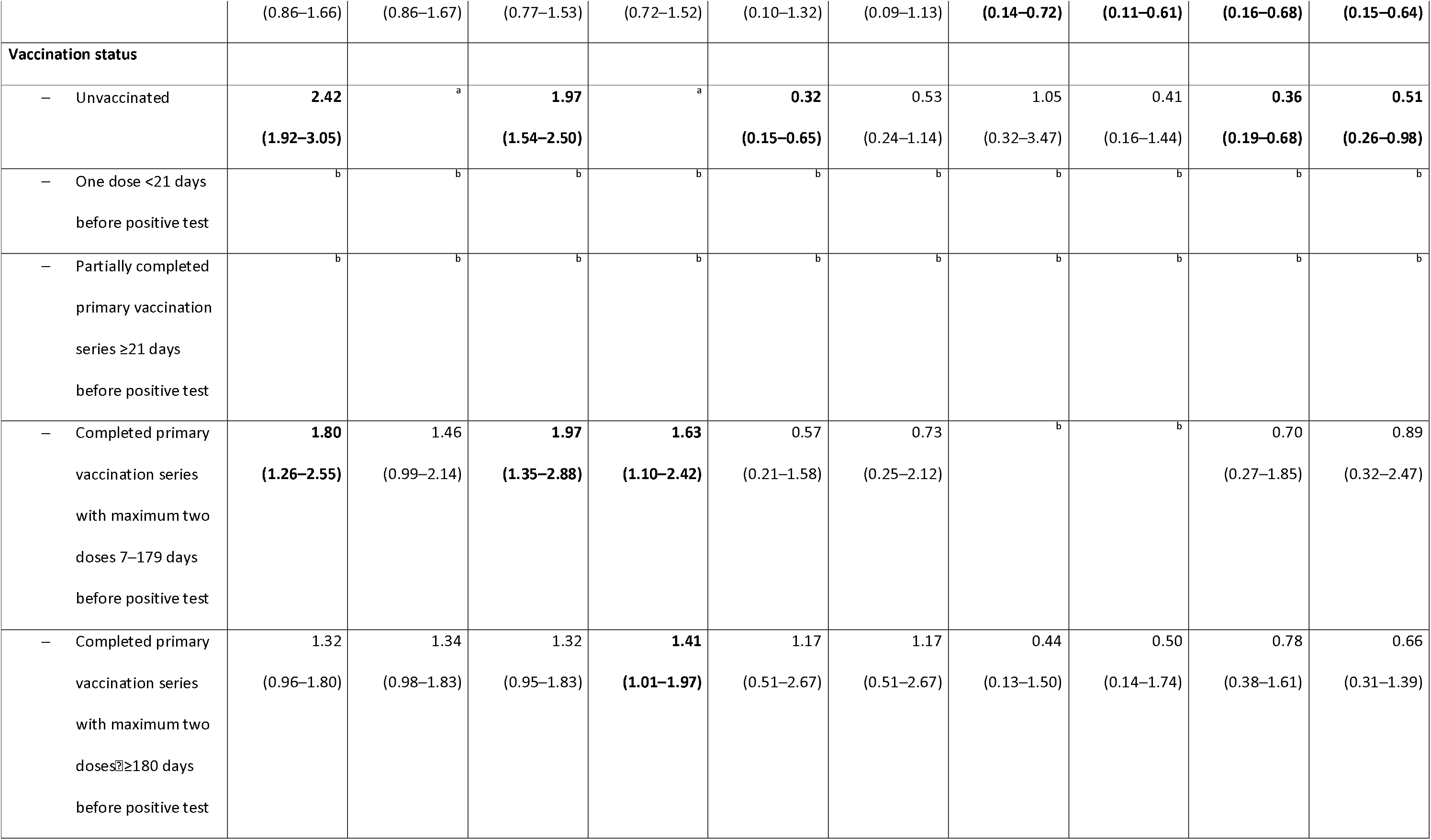
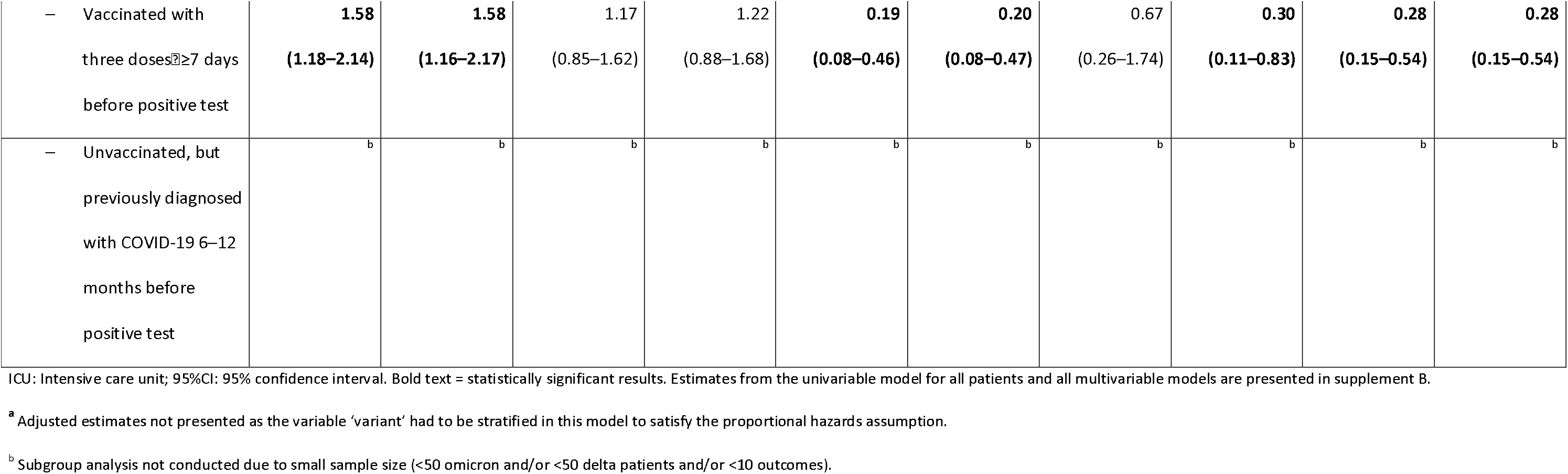
Crude and adjusted hazard ratios for discharge from hospital with and without stay in intensive care, intensive care admission and/or in-hospital death from a Cox proportional hazards model, patients admitted to hospital with omicron variant compared with patients admitted with delta variant of SARS-CoV-2, overall and by subgroup, Norway, 6 December 2021 – 6 February 2022

In the multivariable model including all patients, the variable ‘variant’ did not satisfy the proportional hazards assumption for either LoS outcome. However, our subgroup analysis suggested a shorter LoS (with or without ICU stay) for Omicron patients compared to Delta patients in the age subgroups 18– 79 years and those who had completed primary vaccination. For example, for Omicron patients vaccinated with three doses the aHR for discharge overall was 1.58 (95%CI: 1.16–2.17). Assuming exponential distribution of the survival data (see supplement A, part 2.3), this translates into an expected 37% (95%CI: 14%–54%) shorter overall LoS for Omicron patients (1-(1/1.58)) (table 3).

## Discussion

We find that hospitalised COVID-19 patients infected with the Omicron variant have a milder disease trajectory than patients infected with Delta in Norway. This supports the growing evidence of reduced disease severity among those infected with Omicron (2;4-8). Results from similar studies in South Africa, USA and France have also reported a reduction in the median LoS, risk of ICU admission and/or death among Omicron patients compared to Delta patients (9-12).

Our subgroup analyses generally supported the main results, although we did not observe any difference in the risk of ICU admission or death between Omicron and Delta patients who had completed primary vaccination with maximum two doses. This may be in line with evidence of reduced vaccine effectiveness against infection with Omicron (13-15), and in Norway we have previously observed similar results when analysing the risk of hospitalisation among reported COVID-19 cases (2). However, we did not observe an interaction between variant and vaccination status in this study, while the size of each subgroup must be considered. Such relationships should be further explored in larger patient cohorts.

We have analysed national data from a cohort of patients with known variant data, encompassing 63% of all hospitalisations due to COVID-19 in the study period. One limitation with our study is that a higher proportion of patients admitted to ICU had known variant (see supplement A, part 3). Given the increased risk of ICU admission for Delta patients, we may have oversampled severely ill Delta patients, which may cause us to slightly overestimate the reduction in our outcomes for Omicron patients. Another important limitation is that we could not distinguish sublineage BA.1 and BA.2 for all Omicron patients. In Norway BA.2 has gradually begun to outcompete BA.1 (16). However, up to the end of the study period, BA.1 was still the dominant circulating Omicron sublineage, and our results were robust when we excluded 57 patients known to be infected with BA.2 (see supplement A, part 2.4). Further studies are needed to investigate whether disease severity differs between Omicron sublineages.

Evidence of lower disease severity among hospitalised Omicron patients in Norway and elsewhere is encouraging in the ongoing response to the COVID-19 pandemic. Analyses of circulating variants in a local context are important for informing decision-making on control measures and hospital capacity planning in different settings.

## Data Availability

All data produced in the present study are available upon reasonable request to the authors.

## Notes and acknowledgements

### Authors’ contributions

All co-authors were involved in the conceptualisation of the study. RW coordinated the study. OH, RK, KB and EB contributed to the acquisition of data. LV, HB, JS, OH, NA, ES, KB and RW contributed to data cleaning, verification and/or preparation. ABK, JS, ES, HB, LV and RW had access to the final linked dataset. ABK conducted the statistical analysis in consultation with JS and RW. JS and RW drafted the manuscript. All co-authors contributed to the interpretation of the results. All co-authors contributed to the revision of the manuscript and approved the final version for submission.

### Conflict of interest

The authors declare that they have no competing interests.

### Funding

This research did not receive any specific grant from funding agencies in the public, commercial, or not-for-profit sectors.

### Data sharing statement

The dataset analysed in the study contains individual-level linked data from various central health registries, national clinical registries and other national administrative registries in Norway. The researchers had access to the data through the national emergency preparedness registry for COVID-19 (Beredt C19), housed at the Norwegian Institute of Public Health (NIPH). In Beredt C19, only fully anonymised data (i.e. data that are neither directly nor potentially indirectly identifiable) are permitted to be shared publicly. Legal restrictions therefore prevent the researchers from publicly sharing the dataset used in the study that would enable others to replicate the study findings. However, external researchers are freely able to request access to linked data from the same registries from outside the structure of Beredt C19, as per normal procedure for conducting health research on registry data in Norway. Further information on Beredt C19, including contact information for the Beredt C19 project manager, and information on access to data from each individual data source, is available at https://www.fhi.no/en/id/infectious-diseases/coronavirus/emergency-preparedness-register-for-covid-19/.

## Acknowledgements

First and foremost, we wish to thank all those who have helped report data to the national emergency preparedness registry at the Norwegian Institute of Public Health (NIPH) throughout the pandemic. We also highly acknowledge the efforts that regional laboratories have put into establishing a routine variant screening procedure or whole genome sequencing at short notice and registration of all analysis in national registries for surveillance. Thanks also to the staff at the Virology and Bacteriology departments at NIPH involved in national variant identification and whole genome analysis of SARS-CoV-2 viruses. We also highly acknowledge the efforts of staff at hospitals around Norway to ensure the reporting of timely and complete data to the Norwegian Intensive Care and Pandemic Registry, as well as colleagues at the register itself. We would also like to thank Anja Elsrud Schou Lindman, project director for the national preparedness registry, and all those who have enabled data transfer to this registry, especially Gutorm Høgåsen at the NIPH, who has been in charge of the establishment and administration of the registry.

## Notes

### Competing Interest Statement

The authors have declared no competing interest.

### Funding Statement

The authors received no specific funding for this work.

### Author Declarations

Ethical approval was granted by Regional Committees for Medical Research Ethics, Southeast-Norway, reference number 249509.

