## Supplementary material A for "Milder disease trajectory among COVID-19 patients hospitalised with the SARS-CoV-2 Omicron variant compared with the Delta variant in Norway"

**Supplement A: Shorter length of hospital stay and reduced risk of intensive care admission and in-hospital death among COVID-19 patients hospitalised with the SARS-CoV-2 Omicron variant compared with the Delta variant in Norway, December 2021 – February 2022**

**1. Additional information on the data sources and definitions**

All data in this study came from the [national emergency preparedness register](https://www.fhi.no/en/id/infectious-diseases/coronavirus/emergency-preparedness-register-for-covid-19/), Beredt C19. Beredt C19 contains individual-level data from central health registries, national clinical registries and other national administrative registries in Norway.

*1.1 Admission to hospital and an intensive care unit*

We obtained data on hospitalisation with a positive SARS-CoV-2 test from the Norwegian Intensive Care and Pandemic Registry (NIPaR). All Norwegian hospitals report to NIPaR, and reporting is mandatory. Hospitals in Norway functioned within capacity during the study period, and criteria for hospitalisation and isolation for COVID-19 patients were consistent.

For patients who contracted COVID-19 while admitted to hospital, the time of admission is set to the date of symptom onset, or date of sampling if the patient is asymptomatic. The time of discharge for patients who remain in hospital after they recover from COVID-19 is set to the date they recovered from COVID-19, or if asymptomatic the date that the patient comes out of isolation. The reported main cause of hospitalisation is a clinical assessment. For patients reported with a different main cause than COVID-19, we cannot rule out that COVID-19 may have been a contributing factor for admission. There is no reason to believe, however, that this assessment would differ between patients infected with different variants.

In NIPaR, underlying risk factors for severe COVID-19 diagnosed before admission are registered. The following risk factors are registered; asthma, cancer, chronic lung disease, chronic neurological or neuromuscular disease, diabetes (type 1 and 2), heart disease including hypertension, immunocompromised including HIV and immunosuppressive treatment (includes ongoing use of steroids in doses equivalent to at least 5mg Prednisolone daily), kidney disease including kidney failure, liver disease including liver failure, pregnancy, current smoker and body mass index (calculated as weight in kilograms divided by height in metres squared). For cancer, only active cancer is registered, meaning cancer where the patient still receives treatment, or regular control (>1 per year). Other well-regulated or treated conditions are not distinguished from unregulated or untreated conditions, for example asthma. Full details on the registration of hospitalised patients are available here (in Norwegian): <https://helse-bergen.no/norsk-pandemiregister/registrering-i-norsk-pandemiregister-informasjon-til-ansatte>

NIPaR also includes data on patients who have tested positive for COVID-19 and are admitted to an intensive care unit (ICU). Patients are registered as ICU patients if they fulfil one of five categories:

1. Length of stay over 24 hours in intensive care
2. Require mechanical ventilation
3. Are transferred between intensive care wards
4. Persistent administration of vasoactive medication
5. Length of stay under 24 hours, but passed away during stay in intensive care

Full details on the registration of ICU patients are available here (in Norwegian): <https://helse-bergen.no/norsk-pandemiregister/registrering-i-norsk-pandemiregister-informasjon-til-ansatte>. ,

*1.2 Laboratory testing for variants*

Data on virus variants came from the MSIS laboratory database (Norwegian Surveillance System for Communicable Diseases’ laboratory database), which receives SARS-CoV-2 test results from all Norwegian microbiology laboratories. Variants are identified based on whole genome sequencing, Sanger partial S-gene sequencing or PCR screening targeting specific single nucleotide polymorphisms, insertions or deletions that reliably differentiate between omicron and other variants. The laboratory testing for variants of SARS-CoV-2 in Norway has been described in further detail at: <https://www.fhi.no/nettpub/coronavirus/testing/pavisning-og-overvakning-av-sars-cov-2-virusvarianter/>.

*1.3 National identity number*

Data on persons with a national identity number was drawn from the national population registry. The national identity number was essential to link data from all registries used in the analysis. Data from the population registry was also used to categorise patients by country of birth.

*1.4 Vaccination status and vaccine type*

We defined patients according to the number of vaccine doses received and previously diagnosed COVID-19 cases at date of positive test. Data on COVID-19 vaccinations came from the Norwegian Immunisation Registry, SYSVAK. Data on date of positive test and previous laboratory-confirmed cases of COVID-19 came from the Norwegian Surveillance System for Communicable Diseases (MSIS). As of February 2022, previous laboratory-confirmed cases of COVID-19 are registered in MSIS if there are ≥6 months between two positive sampling dates for an individual. This will exclude reinfections within a 6-month period, of which the Omicron variant could be of higher risk. We also cannot rule out that there were other previously undiagnosed SARS-CoV-2 infections in our cohort, regardless of time between infections.

Categorisation of patients by vaccination status:

1.	Unvaccinated with a COVID-19 vaccine before positive test. Patients who were unvaccinated but had been previously diagnosed with COVID-19 6–12 months before positive test were categorised separately.

2.	Vaccinated with 1 dose of a COVID-19 vaccine <21 days before positive test.

3.	Partially completed primary vaccination series ≥21 days before positive test – those who tested positive ≥21 days after their first dose of a COVID-19 vaccine with a minimum two-dose primary series, and <7 days after the second dose.

4.	Completed primary vaccination series with maximum two doses before positive test – those who tested positive ≥7 days after their second dose and <7 days after their third dose, with at least the recommended minimum interval between doses depending on the type of vaccine (<https://www.fhi.no/om/koronasertifikat/til-helsepersonell-vanlige-problemstillinger-om-koronasertifikat/#oversikt-over-intervall-mellom-koronavaksiner>). This group also includes persons who had tested positive ≥7 days after their first vaccine dose if they had previously also been diagnosed with COVID-19 ≥21 days before or after vaccination and 6–12 months before their current positive test (n=3 for Omicron patients, 2.1% of all Omicron patients with two doses; n=0 for Delta patients). Patients who received the Janssen vaccine® (Janssen Vaccines, Leiden, Netherlands) were also included in this category if they tested positive ≥21 days after one dose (n=0 for Omicron patients; n=2 for Delta patients, 1.2%). We further divided up this two-dose category into those who had received their last dose 7–179 days before positive test, and those who had received their last dose ≥180 days before positive test. Among those who had received their last dose ≥180 days before positive test the median time from last dose to date of positive test was 239 days (interquartile range (IQR): 207–256) for Omicron patients (n=69) and 210 days (IQR: 194–246) for Delta patients (n=111).

5.	Three doses before positive test – those who tested positive ≥7 days after their third dose. This category predominantly includes persons who received their third dose as a booster dose, however it will also include patients who received their third dose as part of their primary series, for example those severely immunocompromised (see <https://www.fhi.no/nyheter/2021/flere-med-alvorlig-nedsatt-immunforsvar-bor-ta-3.-dose-koronavaksine/>). We were not able to clearly distinguish persons who had received a third dose as part of their primary series from those who had received a booster dose. Nine patients (8 Omicron and 1 Delta) had received four vaccine doses, meaning a booster dose following a three-dose primary series. This category also includes persons who had tested positive ≥7 days after their second vaccine dose if they had previously also been diagnosed with COVID-19 ≥21 days before or after vaccination and 6–12 months before their current positive test (n=5 for Omicron patients, 3.3% of all Omicron patients with three doses; n=0 for Delta patients). Among those who had received three doses before positive test the median time from last dose to date of positive test was 68.5 days (interquartile range (IQR): 45–95.5) for Omicron patients (n=152) and 44 days (IQR: 21–77) for Delta patients (n=71).

In Norway, the mRNA vaccines Comirnaty® (BioNTech-Pfizer, Mainz, Germany/New York, United States) and Spikevax® (mRNA-1273, Moderna, Cambridge, United States) have been the two predominant vaccines administered. Among the 298 Omicron patients who had completed their primary vaccination series, 215 (72%) had received a homologous Comirnaty regimen, 53 (18%) had received a mix of Comirnaty and Spikevax and 27 (9.1%) had received a homologous Spikevax regimen. The remaining 3 (1.0%) had received a mix of Comirnaty or Spikevax with Vaxzevria® (AstraZeneca, Cambridge, United Kingdom), or a homologous Vaxzevria regimen. Among the 245 Delta patients who had completed their primary vaccination series, 198 (81%) had received a homologous Comirnaty regimen, 25 (10%) had received a mix of Comirnaty and Spikevax and 20 (8.2%) had received a homologous Spikevax regimen. The remaining 2 (0.8%) had received Janssen.

### 2. Statistical analysis

*2.1 Outcome measures*

Our outcomes were discharge from hospital (with and without ICU stay), admission to ICU, in-hospital death and a composite measure of ICU admission or in-hospital death. We calculated length of stay (LoS) as the time between first admission and last discharge. Patients with unknown date of discharge from their last stay were considered to still be hospitalised. In-hospital death was registered at discharge.

For patients with several registered ‘stays’, individual stays were combined into one patient trajectory per person so that each individual patient is included once. Separate stays may have been registered if a patient was discharged and readmitted, or transferred between wards or hospitals. For patients with >1 registered hospital stay, we included the time between consecutive stays in the calculation of LoS if <24 hours. Of the 1075 patients included in the study, 48 were readmitted (>24 hours between the two registered stays), 15 (31%) of these were Omicron and 33 (69%) were Delta. Twenty-four Delta patients and nine Omicron patients were transferred between hospitals.

*2.2 Statistical analysis*

Explanatory variables to analyse differences in our outcomes were virus variant, sex, age, country of birth, underlying risk factors, regional health authority and vaccination status. See section 1 and table 1 in the main manuscript for the categorisation of explanatory variables.

Outcomes were explored univariably in a Cox proportional hazards model and by calculating Kaplan Meier curves, with right censoring of patients still admitted to hospital. Crude log hazard ratios with medians and IQR for LoS were obtained. Explanatory variables with p<0.2 were further explored in multivariable models. Forward model selection was performed based on Akaike Information Criterion. Only variables with correlation <0.5 were used in the same model. Virus variant was maintained in all models regardless of significance. Age was tested linearly, with a spline or categorically. The multivariable model was checked for the assumption of proportional hazard by checking Schoenfeld residuals, and explanatory variables were stratified to satisfy the assumption as necessary. We also checked for interactions between variables included in multivariable models. We did not observe an interaction between variant and age or variant and vaccination status in this analysis. Adjusted log hazard ratios (aHR) obtained in the multivariable models were reported. We also conducted subgroup analysis by age group and vaccination status for subgroups with ≥50 omicron and ≥50 delta patients, and ≥10 outcomes.

For LoS outcomes, as hazard rates are not explicitly estimated in Cox regression, we also estimated a proxy for the expected difference in LoS as 1-(1/aHR), by assuming an exponential survival distribution. The fit of LoS outcomes to an exponential distribution is presented in section 2.3.

Estimates from the univariable model for all patients and all multivariable models are presented in supplement B. The statistical analysis was performed in R version 3.6.2.

*2.3 Fit of LoS outcomes to an exponential distribution*

For LoS, as hazard rates are not explicitly estimated in Cox regression, we estimated a proxy for the expected difference in LoS as 1-(1/aHR), by assuming a constant baseline hazard rate, i.e. an exponential survival distribution^[1]^. Figure S1 shows the fit of LoS data to an exponential distribution by age and vaccination status subgroup. Only distributions for statistically significant LoS outcomes are presented (see table 3 in the manuscript).

Figure S1 shows that all observed LoS distributions fit an exponential distribution relatively well for the vast majority of patients up to a given LoS. For patients aged 18–44 years, the observed overall LoS was generally longer than expected for a LoS ≥20 days (see panel A), which corresponds to 14 (5.6%) of patients in this subgroup, of whom 13 (93%) were infected with Delta. For patients aged 45–64 years, the observed overall LoS was generally longer than expected for a LoS ≥25 days (see panel C), which corresponds to 33 (9.6%) of patients in this subgroup, of whom 32 (97%) were infected with Delta. This will cause us to underestimate the percent decrease in LoS for Omicron patients compared to Delta patients using the formula 1-(1/aHR). Observed distributions for LoS for all patients who had been vaccinated with three doses and for patients without admission to intensive care regardless of subgroup did fit an exponential distribution well, aside from a handful of outliers (8 or fewer, predominantly Delta patients) who had a longer LoS than expected.

A)
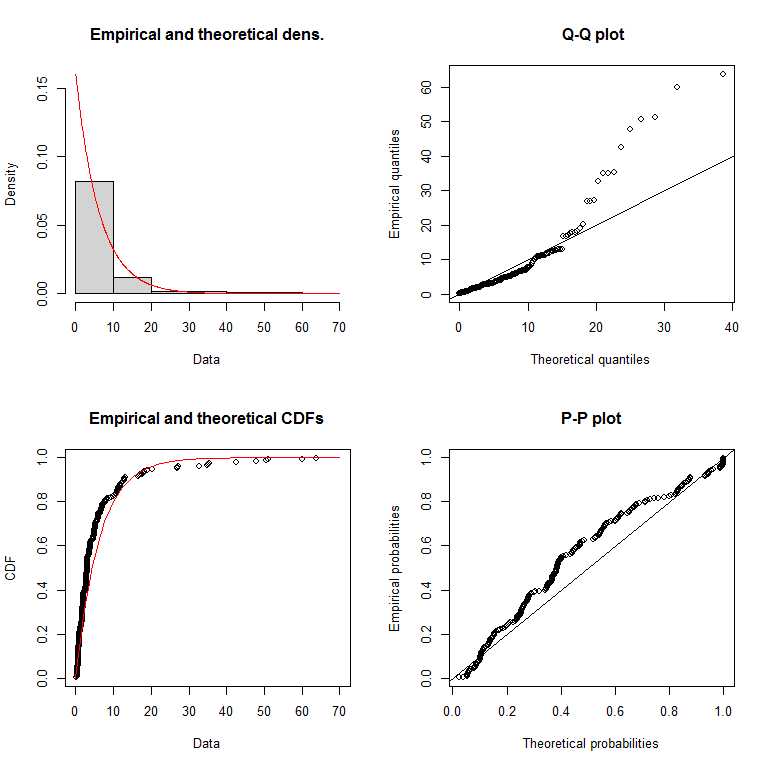
 B)
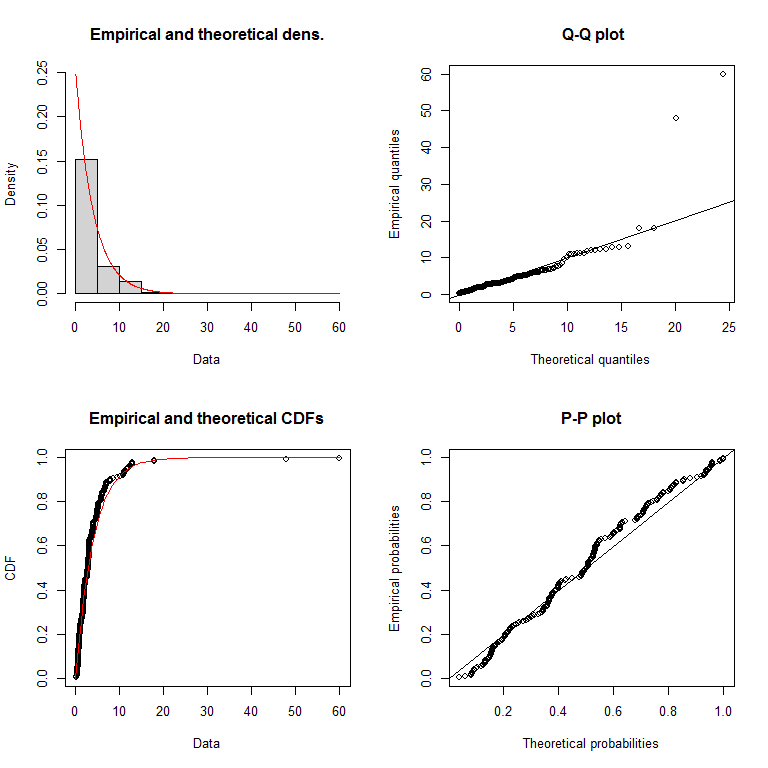

C)
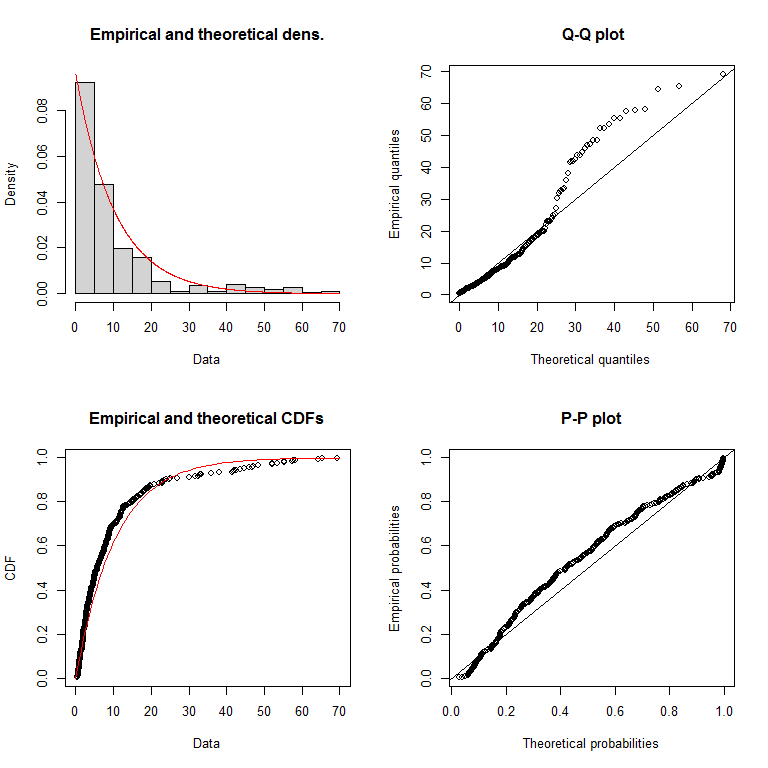
D)
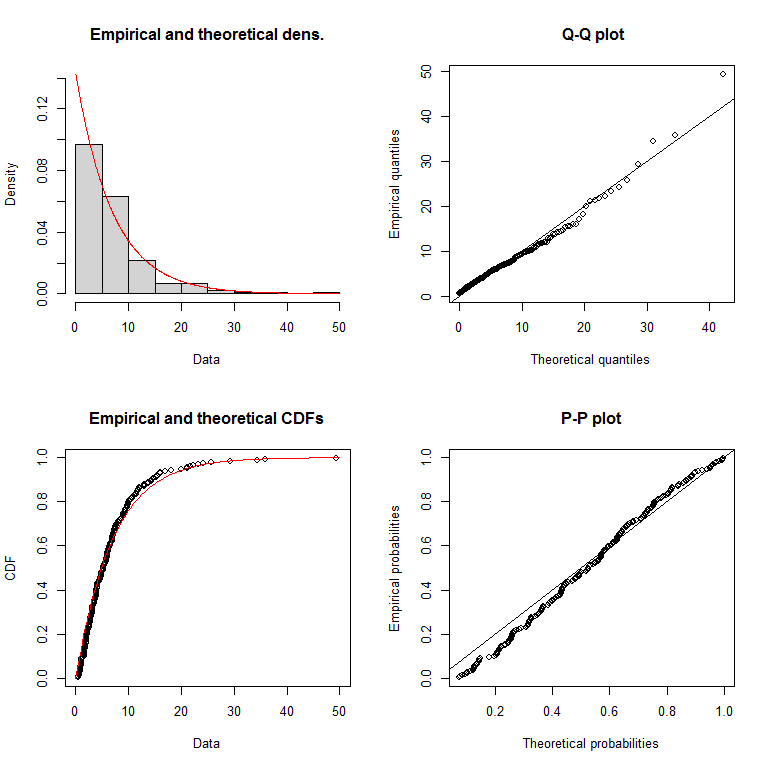

E)
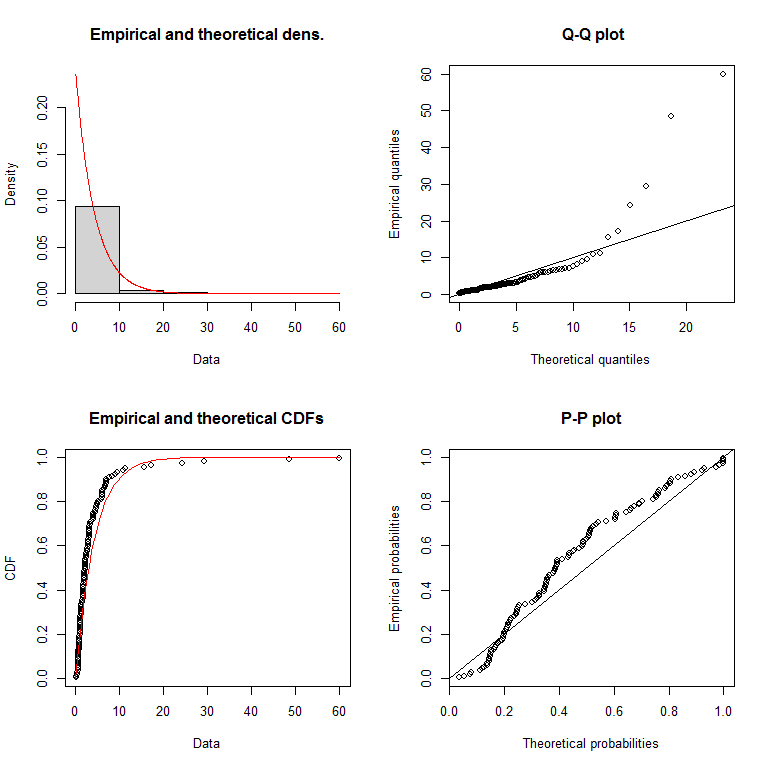
F)
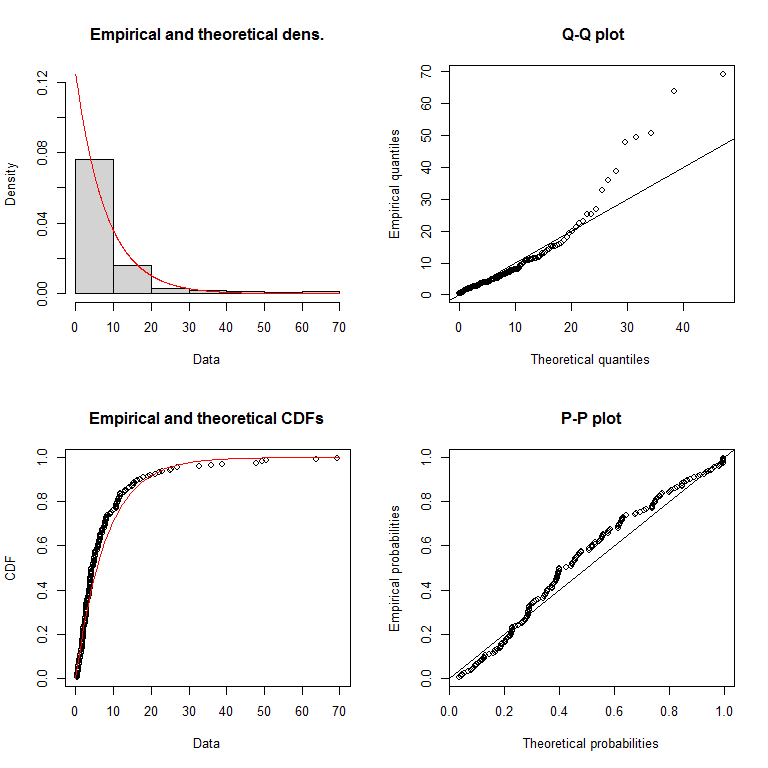

G)
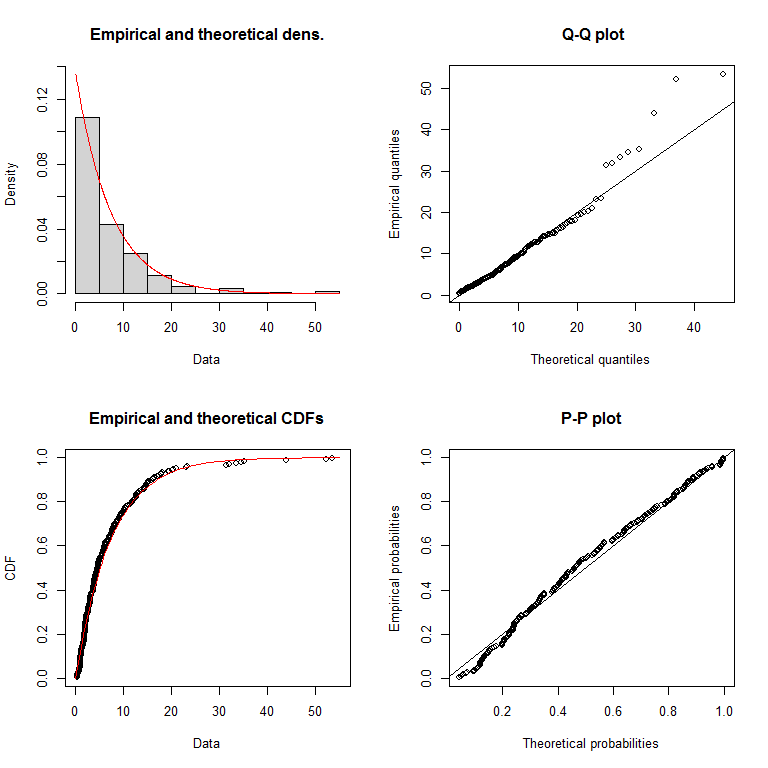

*Figure S1. Fit of length of stay in hospital to an exponential distribution. A) all patients 18–44 years, B) patients 18–44 years not admitted to intensive care, C) all patients 45–64 years, D) patients 65–79 years not admitted to intensive care, E) patients not admitted to intensive care who had completed primary vaccination series with maximum two doses 7–179 days before positive test F) patients not admitted to intensive care who had completed primary vaccination series with maximum two doses ≥180 days before positive test, G) all patients who had been vaccinated with three doses ≥7 days before positive test.*

*2.4 Sensitivity analysis excluding patients known to be infected with Omicron subvariant BA.2.*

The Omicron variant is considered to comprise all viruses belonging to the Pango lineage B.1.1.529. This lineage is subdivided into distinct sublineages such as BA.1 (alias for B.1.1.529.1) and BA.2 (alias for B.1.1.529.2). BA.1 superseded Delta as the dominant circulating strain by the end of December. Between week 1 and 5 2022, over 90 % of the notified cases has been infected with the Omicron BA.1 variant. From week 5, the prevalence of Omicron BA.1 decreased, while the prevalence of Omicron BA.2 increased. By week 7, 19 % of sequenced cases were Omicron BA.2^[2]^.

As it was not possible for us to distinguish Omicron sublineages for all patients infected with Omicron, it was not possible for us to do a separate analysis comparing BA.1 and BA.2. However, in addition to the main analysis, we conducted a sensitivity analysis excluding the 57 patients known to be infected with BA.2, to further explore the impact of these cases on the main analysis. The methodology was the same as used in the main analysis. As shown in Table S1, estimates for all outcomes were robust.

*Table S1. Sensitivity analyses excluding patients known to be infected with Omicron subvariant BA.2 for intensive care admission, in-hospital death and intensive care admission or in-hospital death from a Cox proportional hazards model, by virus variant, Norway, 6 December 2021 – 6 February 2022.*

| Variant | Number of patients | Admission to ICU | | Death in hospital | | ICU admission OR death in hospital | |
| --- | --- | --- | --- | --- | --- | --- | --- |
|  |  | Yes | Adjusted hazard ratio (95%CI) | Yes | Adjusted hazard ratio (95%CI) | Yes | Adjusted hazard ratio (95%CI) |
| *Main analysis* | | | | | | | |
| Delta | 666 | 165 | Ref | 63 | Ref | 206 | Ref |
| Omicron | 409 | 31 | **0.52**  **(0.34–0.80)** | 15 | **0.44**  **(0.24–0.79)** | 42 | **0.46**  **(0.32–0.67)** |
| *Excluding 57 patients known to be infected with subvariant BA.2 from the Omicron cohort* | | | | | | | |
| Delta | 666 | 165 | Ref | 63 | Ref | 206 | Ref |
| Omicron | 352 | 29 | **0.55**  **(0.36–0.85)** | 14 | **0.46**  **(0.25–0.83)** | 39 | **0.49**  **(0.33–0.71)** |

ICU: Intensive care unit; 95%CI: 95% confidence interval. Bold text = statistically significant results. Length of stay outcomes not included as the variable ‘variant’ had to be stratified in the model with all patients for these outcomes to satisfy the proportional hazards assumption.

### 3. Assessment of representativeness of study population

We assessed the representativeness of our study population by comparing the characteristics of patients who had data on the SARS-CoV-2 variant that they were infected with, with those who did not. Of 1710 patients diagnosed in the study period with a national identity number, 1079 (63%) were screened. We found differences between cases who were screened for variants with regards to week of admission, regional health authority, age, vaccination status, country of birth, some risk factors (chronic neurological or neuromuscular disease, immunosuppression, kidney disease) and admission to intensive care (Table S2).

The proportion of screened patients among patients admitted to ICU was larger than those who were not admitted to ICU (79% vs 60%). Therefore, our cohort of patients might be more representative of severely ill patients. This may also explain a higher proportion of screened patients in certain age groups (e.g. 65-79 years: 71%) compared to others (e.g. 0-17 years: 51%), and a higher proportion of screened patients with certain underlying medical conditions that give a higher risk of a severe disease course, such as chronic neurological or neuromuscular disease, immunosuppression and kidney disease.

Differences among regional health authorities were also observed, with a lower proportion of patients in the North regional health authority screened compared to the other health authorities. This reflects a difference in screening capacity in the different health authorities. Further, more cases were screened during the first weeks of the study period compared to the last weeks because screening activity was reduced after Omicron became the dominant circulating variant.

*Table S2: Characteristics of reported cases by whether or not they were screened for the SARS-CoV-2 variant that they were infected with, 6 December 2021 – 6 February 2022, Norway.*

| Characteristics | | Known variant | | |
| --- | --- | --- | --- | --- |
|  |  | No (n=631) | Yes (n=1079) | p value |
| Sex | Male | 331 (36.1%) | 586 (63.9%) | 0.458 |
|  | Female | 300 (37.8%) | 493 (62.2%) |  |
| Age group | 0-17 years | 57 (49.1%) | 59 (50.9%) | <0.001 |
|  | 18-29 years | 61 (51.3%) | 58 (48.7%) |  |
|  | 30-44 years | 112 (36.8%) | 192 (63.2%) |  |
|  | 45-54 years | 93 (36.8%) | 160 (63.2%) |  |
|  | 55-64 years | 96 (34.4%) | 183 (65.6%) |  |
|  | 65-79 years | 113 (29.5%) | 270 (70.5%) |  |
|  | ≥80 years | 99 (38.7%) | 157 (61.3%) |  |
| Median age | In years (IQR) | 54 (35–73) | 58 (42–74) | <0.001 |
| Born in Norway | Yes, with at least one parent born in Norway | 331 (34.4%) | 631 (65.6%) | <0.001 |
|  | Yes, two parents born outside of Norway | 20 (37.7%) | 33 (62.3%) |  |
|  | No | 207 (36.8%) | 356 (63.2%) |  |
|  | Unknown | 73 (55.3%) | 59 (44.7%) |  |
| Asthma | Yes | 59 (35.1%) | 109 (64.9%) | 0.614 |
|  | No | 572 (37.1%) | 970 (62.9%) |  |
| Cancer ^a^ | Yes | 40 (32.0%) | 85 (68.0%) | 0.238 |
|  | No | 591 (37.3%) | 994 (62.7%) |  |
| Chronic lung disease, excluding asthma | Yes | 70 (33.2%) | 141 (66.8%) | 0.231 |
|  | No | 561 (37.4%) | 938 (62.6%) |  |
| Chronic neurological or neuromuscular disease | Yes | 36 (27.7%) | 94 (72.3%) | 0.023 |
|  | No | 595 (37.7%) | 985 (62.3%) |  |
| Diabetes (type 1 and 2) | Yes | 85 (33.2%) | 171 (66.8%) | 0.184 |
|  | No | 546 (37.6%) | 908 (62.4%) |  |
| Heart disease, including hypertension | Yes | 200 (34.4%) | 382 (65.6%) | 0.118 |
|  | No | 431 (38.2%) | 697 (61.8%) |  |
| Immunosuppression, including HIV and immunosuppressive treatment ^b^ | Yes | 51 (29.5%) | 122 (70.5%) | 0.033 |
|  | No | 580 (37.7%) | 957 (62.3%) |  |
| Kidney disease, including kidney failure | Yes | 52 (29.2%) | 126 (70.8%) | 0.025 |
|  | No | 579 (37.8%) | 953 (62.2%) |  |
| Liver disease, including liver failure | Yes | 9 (29.0%) | 22 (71.0%) | 0.360 |
|  | No | 622 (37.0%) | 1057 (63.0%) |  |
| BMI ≥30 | Yes | 95 (32.1%) | 201 (67.9%) | 0.102 |
|  | No | 240 (37.6%) | 398 (62.4%) |  |
|  | Unknown | 296 (38.1%) | 480 (61.9%) |  |
| Pregnant | Yes | 26 (43.3%) | 34 (56.7%) | 0.293 |
|  | No | 605 (36.7%) | 1045 (63.3%) |  |
| Current smoker | Yes | 25 (28.4%) | 63 (71.6%) | 0.090 |
|  | No | 606 (37.4%) | 1016 (62.6%) |  |
| Vaccination status | Not vaccinated | 281 (35.9%) | 501 (64.1%) | 0.004 |
|  | One dose <21 days before positive test | 8 (38.1%) | 13 (61.9%) |  |
|  | Partially completed primary vaccination series ≥21 days before positive test | 29 (61.7%) | 18 (38.3%) |  |
|  | Completed primary vaccination series with maximum two doses 7–179 days before positive test | 78 (35.6%) | 141 (64.4%) |  |
|  | Completed primary vaccination series with maximum two doses ≥180 days before positive test | 77 (29.4%) | 185 (70.6%) |  |
|  | Vaccinated with three doses ≥7 days before positive test | 131 (37.3%) | 220 (62.7%) |  |
|  | Unvaccinated, but previously diagnosed with COVID-19 6–12 months before positive test | 0 (0.0%) | 1 (100.0%) |  |
|  | Unknown vaccination status | 27 (100.0%) | 0 (0.0%) |  |
| Week of admission | 49/2021 | 56 (25.2%) | 166 (74.8%) | <0.001 |
|  | 50/2021 | 73 (29.9%) | 171 (70.1%) |  |
|  | 51/2021 | 60 (30.3%) | 138 (69.7%) |  |
|  | 52/2021 | 57 (30.6%) | 129 (69.4%) |  |
|  | 1/2022 | 31 (21.8%) | 111 (78.2%) |  |
|  | 2/2022 | 30 (25.4%) | 88 (74.6%) |  |
|  | 3/2022 | 78 (53.8%) | 67 (46.2%) |  |
|  | 4/2022 | 118 (54.4%) | 99 (45.6%) |  |
|  | 5/2022 | 128 (53.8%) | 110 (46.2%) |  |
| Regional health authority | South-East | 412 (36.7%) | 712 (63.3%) | <0.001 |
|  | West | 114 (35.4%) | 208 (64.6%) |  |
|  | Mid | 48 (30.2%) | 111 (69.8%) |  |
|  | North | 57 (54.3%) | 48 (45.7%) |  |
| Admission to ICU | No | 580 (39.6%) | 883 (60.4%) | <0.001 |
|  | Yes | 51 (20.6%) | 196 (79.4%) |  |

IQR: interquartile range; ICU: Intensive care unit; BMI: Body mass index. P values comparing patients with omicron and delta calculated using chi-squared tests or Wilcoxon rank sum tests as appropriate.

^a^ Refers to cancer patients undergoing treatment or with regular controls (>1 per year).

^b^ Includes ongoing use of steroids in doses equivalent to at least 5mg Prednisolone daily.

1. Barraclough H, Simms L, Govindan R. Biostatistics primer: what a clinician ought to know: hazard ratios. J Thorac Oncol. 2011;6(6):978-82
2. Folkehelseinstituttet 2022: COVID-19 ukesrapport, uke 7: <https://www.fhi.no/contentassets/8a971e7b0a3c4a06bdbf381ab52e6157/vedlegg/2022/ukerapport-uke-7-14.02---20.02.22.pdf>
